# Individualized systems-informed corrective exercise for adults meeting prespecified criteria for upper crossed syndrome: protocol for a randomized controlled trial

**DOI:** 10.64898/2026.11.10.26367704

**Authors:** Mohamad Khorami Moghadam, Gholam Ali Ghasemi, Leila PoorSaadat, Masoud Golpayegani

**Affiliations:** Department of Sport Injuries and Corrective Exercises, Faculty of Sport Sciences, University of Isfahan, Isfahan, Iran; Department of Neurology, School of Medicine, Arak University of Medical Sciences, Arak, Iran; Department of Sport Injuries and Corrective Exercises, Faculty of Sport Sciences, Arak University, Arak, Iran

**Keywords:** upper crossed syndrome, corrective exercise, posture, systems perspective, surface electromyography

## Abstract

**Background:** Heterogeneity in eligibility criteria, intervention content, and exercise-progression rules has limited the reproducibility of research on upper crossed syndrome. This trial will examine the added effect of a structured, individualized, staged corrective-exercise program on adults who meet prespecified criteria for this phenotype.

**Methods:** In this parallel randomized controlled trial, 36 women and men aged 18-40 years with forward head and shoulder posture and increased thoracic kyphosis will be allocated 1:1, stratified by sex. Over 12 weeks, both groups will attend three sessions per week and receive stationary cycling, indication-based stretching and mobilization, and education on healthy movement habits. The intervention group will additionally receive a systems-informed program encompassing restoration of range of motion, muscle-activation retraining, correction of movement patterns, progressive load tolerance, and preparation for health-enhancing physical activity. The primary outcome is the baseline-adjusted week-12 between-group difference in a multivariate profile comprising the forward-head, forward-shoulder, and thoracic-kyphosis angles. Scapular-muscle electromyography, range of motion, and balance are secondary outcomes. The primary exploratory electroencephalographic outcome is change in central relative alpha power during quiet standing with eyes open. Posture, range of motion, and balance will be assessed at baseline, week 12, and four-week follow-up; electromyography and electroencephalography at baseline and week 12. The primary analysis will use MANCOVA adjusted for baseline and sex, with Pillai’s trace; longitudinal analyses will use linear mixed models. Effect estimates and 95% confidence intervals will be reported.

**Conclusions:** This study will test the added effect of a reproducible corrective-exercise framework beyond the shared active-control components. The four-week follow-up will not support inference about long-term prevention of recurrence.

**Ethics and trial registration:** Ethics approval: IR.UI.REC.1404.053. Trial registration: IRCT20250921067305N1.

## Introduction

Upper crossed syndrome (UCS) refers to a pattern of postural and neuromuscular changes involving the head, thoracic spine, and shoulder girdle, with forward head posture, forward or rounded shoulders, and increased thoracic kyphosis among its most prominent features. These alterations may occur separately or in different combinations, and previous studies have not used consistent angular thresholds or eligibility criteria to define UCS [1–3]. To avoid making diagnostic claims beyond the available evidence, the present study operationalizes UCS as a postural phenotype requiring the concurrent presence of forward head posture, forward shoulder posture, and increased thoracic kyphosis. This definition is intended to increase sample homogeneity and permit evaluation of a combined postural pattern; it is not equivalent to a validated diagnostic criterion.

Conventional corrective-exercise programs have largely been based on the muscle-imbalance model and have used stretching, strengthening, scapular-stabilization exercises, and postural education [2–7]. A 2024 meta-analysis indicated that therapeutic exercise could improve the three postural components associated with UCS, although substantial heterogeneity existed among studies [2]. A 2026 meta-analysis of 28 trials and 901 participants likewise reported large effects on forward head posture, forward shoulder posture, and thoracic kyphosis, but between-study heterogeneity ranged from 78% to 88% [3]. Pooled findings for muscle activity, balance, range of motion, function, and pain remain uncertain or inconsistent. Thus, the unresolved question is not simply whether exercise can change postural angles, but whether a prespecified and structured framework can improve all three components concurrently and whether these changes are accompanied by neuromuscular responses and maintained after the intervention [2,3]. Although the muscle-imbalance model offers an understandable basis for exercise selection, postural control cannot be reduced to the length, strength, or activity of a small number of muscles [5,6,8].

In this study, a systems perspective provides the theoretical basis for explaining postural and movement control. From this perspective, postural control emerges through continuous interactions among sensory information, central nervous system processing, neuromuscular responses, the body’s mechanical capacities, and the demands of the task and environment, and it adapts to individual and environmental conditions through feedback [9–11]. Drawing on this foundation and on components used in previous comprehensive interventions [6–13], the investigators developed a structured, staged, and individualized framework for the present trial. The intervention is not confined to correcting a single muscle, joint, or postural angle. Instead, modifiable factors within the scope of the intervention are organized into overlapping stages that begin with breathing retraining and trunk control, restoration of mobility, regulation of muscle activation and coordination, and control of movement patterns, and progress toward load tolerance, health-related physical fitness, self-management, and sustained participation in physical activity. Transitions between stages are governed by prespecified criteria relating to individual capacity, movement quality, compensatory movement, and response to loading rather than by elapsed time alone [6,7,9–11]. Accordingly, the novelty of the intervention does not lie in an entirely new exercise or technique, but in how established components are organized, progressed, and transferred [2,3,5,13]. Goals and decision rules are standardized in advance, whereas exercise level is adjusted according to individual capacity, range of motion, execution quality, and compensatory movement. Standardization therefore refers to the use of a shared decision-making framework rather than an identical prescription for every participant. The gradual transition from simple, low-load positions to weight-bearing and daily activities is intended to enhance transfer of learning [14,15].

Findings concerning shoulder-girdle muscle activity reinforce the need to distinguish postural change from neuromuscular response [3,6,7]. Some trials have reported increased serratus anterior activity, reduced upper trapezius activity, or improved scapular-muscle ratios, whereas the available meta-analysis did not confirm consistent changes in selected muscles [3]. Surface electromyography (EMG) is therefore included as an objective secondary outcome, and any change will not be considered definitive evidence of the mechanism of effect.

The contribution of the central nervous system to postural control provides the theoretical rationale for the electroencephalographic (EEG) assessment [9,11,16–18]. Studies of standing and balance learning have reported changes in sensorimotor activity and cortical oscillations, but findings depend on the task, population, and analytical method [16–18]. One study reported EEG differences between forward and neutral head postures [17]; however, an acute change in head position is not equivalent to a UCS-specific brain pattern [18]. Because no UCS-specific EEG biomarker has been established, central relative alpha power at C3, Cz, and C4 during quiet standing with eyes open was prespecified as a single exploratory outcome, based on the sensorimotor relevance of these electrode locations, reported associations between alpha oscillations and postural control, and the lower susceptibility of quiet standing to movement artifacts than more demanding postural tasks. Restricting the analysis to one prespecified band–region–condition combination also reduces multiplicity. EEG is therefore included solely as an exploratory, hypothesis-generating outcome and will not be used as a diagnostic criterion or primary effectiveness outcome. In the absence of source localization, sensor-level findings will not be attributed to specific cortical structures or interpreted as definitive evidence of neural reorganization.

The primary objective of this trial is to determine the added effect of a structured, staged corrective program, compared with a foundational active intervention comprising cycling, stretching, mobilization, and education on healthy movement habits, on the postural profile of adults with the operational UCS phenotype. The primary outcome is the multivariate profile of forward-head, forward-shoulder, and thoracic-kyphosis angles at week 12. The primary hypothesis is that, after adjustment for baseline values, this profile will show greater improvement in the intervention group. EMG, range of motion, and balance are secondary outcomes; maintenance of postural changes at four-week follow-up is a supportive longitudinal analysis; and EEG is exploratory.

## Materials and methods

### Study design and setting

This is a two-arm, parallel-group randomized controlled trial with 1:1 allocation. It is designed to estimate the added effect of a 12-week structured, individualized, staged corrective-exercise program relative to a foundational active intervention. Both groups will receive shared components comprising stationary cycling, indication-based stretching and mobilization, and education on healthy movement habits; only the intervention group will receive the model-specific staged component. The between-group comparison therefore estimates the effect of adding the specific intervention package to the shared components, rather than the effect of the entire intervention compared with no treatment. The unit of randomization is the individual. The protocol was prepared in accordance with SPIRIT 2025 [19,20], the final trial report will follow CONSORT 2025 [21], and the intervention description has been aligned with TIDieR-Rehab and CERT [22,23].

The study received ethics approval from the Research Ethics Committee of the University of Isfahan (IR.UI.REC.1404.053).

The trial was prospectively registered in the Iranian Registry of Clinical Trials on 12 November 2025 (IRCT20250921067305N1; https://en.irct.ir/trial/86313).

Musculoskeletal and balance assessments will be conducted at the Sport Sciences Laboratory of Arak University, EEG recordings at Valiasr Hospital in Arak, and intervention sessions at Diaphragm Clinic in Arak. Screening will occur no more than 14 days before baseline; baseline assessment will occur no more than seven days before randomization; post-intervention assessment will be conducted 48-72 hours after the final session; and follow-up will occur four weeks after the intervention, with an allowable window of ±3 days. The 48-72-hour interval is intended to reduce the influence of acute responses to the final session. The schedule is presented in figure 1.

**Fig 1.**
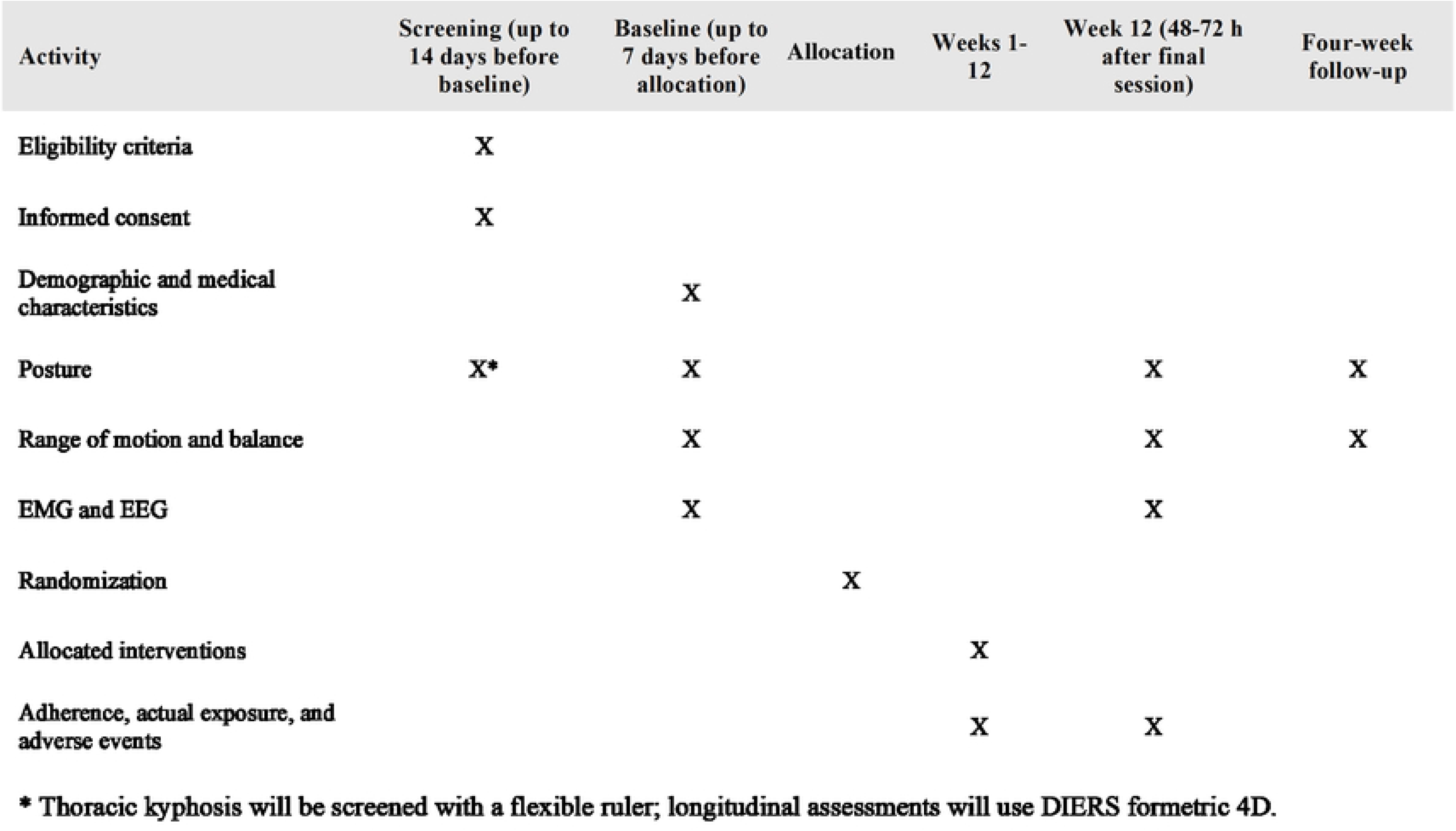
Schedule of enrollment, interventions, and assessments according to SPIRIT

### Trial status

This is protocol version 1.0, dated 12 October 2026. Formal participant recruitment has not yet commenced. Activities to date have been limited to feasibility outreach and preliminary informal screening, during which approximately 10 potentially eligible individuals were identified. No participant has been enrolled or randomized, no intervention has begun, and no outcome data have been collected or analyzed.

Formal recruitment is planned to begin in mid-November 2026 and is expected to be completed by December 2026. Following eligibility screening and baseline assessment, participants will undergo the 12-week intervention and four-week follow-up. All data collection is expected to be completed by April 2027, and the first study results are expected by June 2027. Any substantive deviation from this projected schedule will be documented in the trial registry and final trial report.

### Participants and eligibility criteria

The target population comprises women and men aged 18-40 years with the operational UCS phenotype who live in Arak. Volunteers will be recruited through university advertisements, sports and clinical centers, and social media. Sex will be used as a stratification factor in randomization. Verbal and written study information will be provided, and written informed consent will be obtained before formal assessment. Photographs required for postural measurement will be obtained under the general study consent, with facial identity obscured using a three-layer mask and all files stored under coded identifiers. Separate consent will apply only to identifiable photographs or videos intended for presentation, publication, or dissemination; refusal to provide that additional consent will not preclude participation.

Eligibility for the operational study phenotype requires the concurrent presence of a forward-head angle of at least 44 degrees, a forward-shoulder angle of at least 49 degrees, and thoracic kyphosis of at least 42 degrees. These thresholds were selected before recruitment based on operational cutoffs used in previous UCS protocols and trials; however, in earlier studies, the presence of one of these postural components together with other UCS features was sufficient for inclusion. Requiring all three components in this trial is a prespecified phenotype-enrichment strategy intended to increase sample homogeneity. It is not a validated diagnostic rule, a definitive clinical cutoff, or a comprehensive definition of UCS. Published evidence on reliability and validity pertains to the photogrammetric and flexible-ruler measurement methods and should not be interpreted as diagnostic validation of this three-component combination [7,8,24,25]. The number of individuals excluded because they fail each threshold, and the combinations of failed thresholds will be reported in the study flow diagram. Participants must have a body mass index of 18.5-24.9 kg/m^2^, be able to undertake the tests and intervention safely, and be willing to maintain their usual pattern of activity outside the protocol [26]. The BMI restriction is intended to reduce the influence of subcutaneous tissue on the amplitude and selectivity of surface EMG signals [27].

Exclusion criteria are surgery or fracture affecting the spine, shoulder girdle, or pelvis; a diagnosed neurological, psychiatric, or musculoskeletal condition that could compromise safety or confound the outcomes; medication affecting the central nervous system; acute pain or uncontrolled cardiopulmonary disease; pregnancy; an angle of trunk rotation greater than 5 degrees during the forward-bending test; a lower-limb or pelvic disorder that precludes valid balance testing; and regular participation in corrective exercise, resistance training, or manual therapy during the preceding three months. After randomization, a new illness or injury, a prohibited concurrent intervention, or a medical decision may lead to discontinuation of the assigned intervention; however, the participant will remain in follow-up unless consent is withdrawn. Menstrual-cycle status will be recorded, but cycle irregularity alone will not be an exclusion criterion.

### Randomization, allocation concealment, and blinding

After baseline assessment, an independent statistician will generate the allocation sequence using randomly permuted blocks of four and six, stratified by sex. Block sizes will not be disclosed to personnel involved in recruitment or assessment. Allocations will be placed in sequentially numbered, opaque, tamper-evident, sealed envelopes. An envelope will be opened only after eligibility has been confirmed, baseline assessment completed, and the participant irreversibly enrolled, by a person who has no access to the allocation sequence. Sex will be included in the primary models, but the limited sample precludes subgroup analysis or testing of an intervention-by-sex interaction.

Participants and the intervention provider cannot be blinded. Outcome assessors, EMG and EEG processors, and the statistician will remain blinded to group allocation. Images, Formetric outputs, and signal files will be coded without group or time-point information and presented in random order. Metadata that could reveal group or assessment time will be removed or concealed. The code key will be held by an independent person until completion of the primary analysis, and any inadvertent unblinding will be documented.

### Exercise intervention

The intervention integrates breathing and trunk control, mobility, muscle activation and coordination, segmental alignment, movement quality, and transfer to daily activity within a systems-informed framework. Its operational rationale, exercise bank, decision rules, and fidelity procedures are detailed in S4 File.

The intervention model comprises six numbered, overlapping stages: (1) correction of breathing pattern and core stability; (2) restoration of range of motion; (3) retraining of muscle activation and coordination; (4) basic movements; (5) fundamental movement patterns; and (6) functional movement patterns. These stages are followed by an unnumbered transition toward load tolerance and health-related physical fitness. Maintenance, self-management, and participation in an active lifestyle represent the destination of this continuum and are not confined to a single, discrete session or stage. Stages are not separated by a fixed calendar schedule. Entry into a higher stage is based on individual performance criteria, and exercises from earlier stages may continue to consolidate movement quality. Figure 2 illustrates the overlap between stages and the different progression pathways that participants may follow.

**Fig 2.**
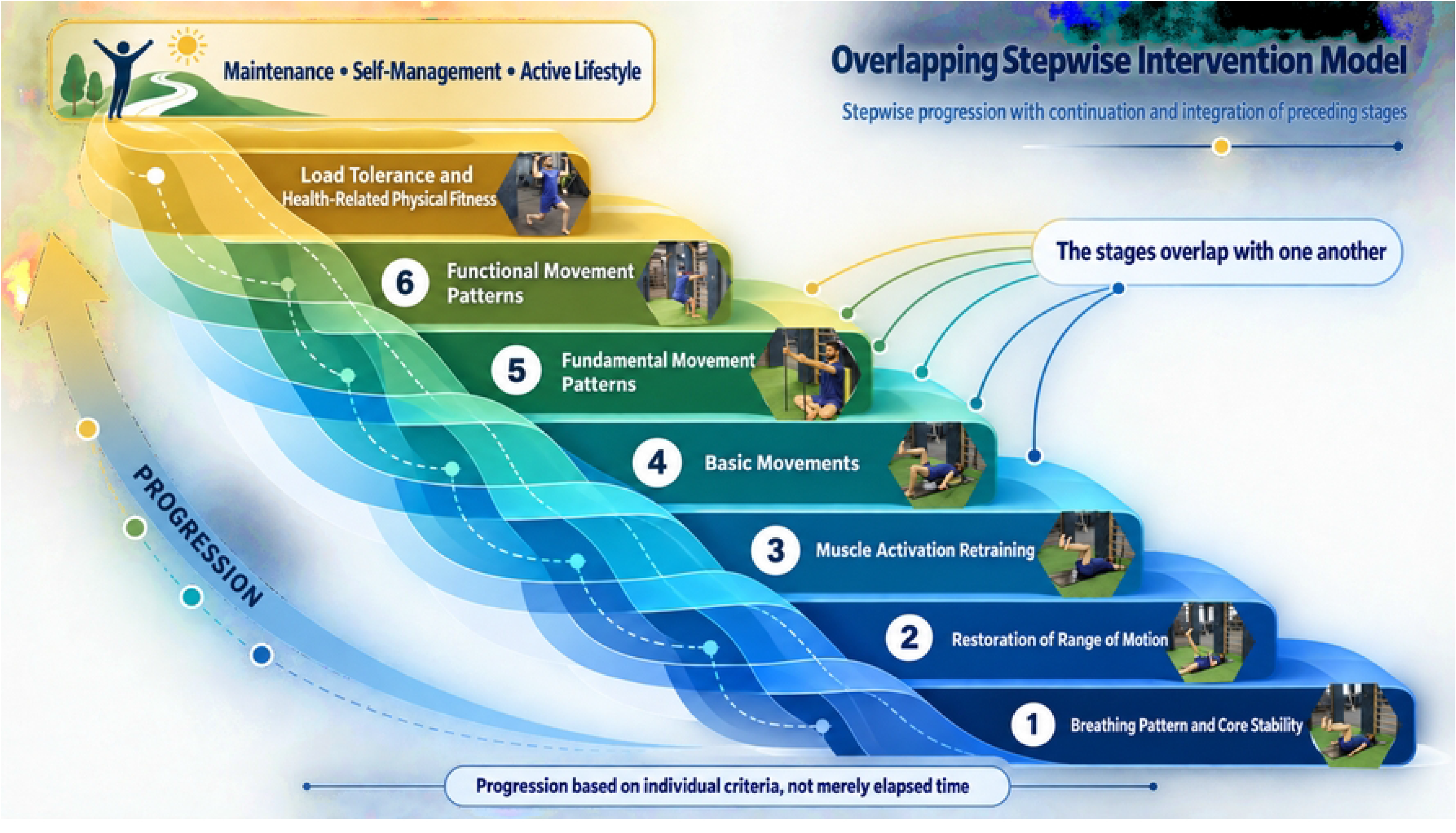
Staged and overlapping model of the structured intervention.

### Delivery and dose

The intervention will be delivered by the principal investigator, a final-year PhD candidate in Sport Injuries and Corrective Exercise who developed the intervention framework and has clinical experience in corrective-exercise assessment, individualized exercise prescription, manual techniques, and exercise progression. Delivery procedures will be standardized according to the supplementary intervention manual before participant enrollment.

The program will be delivered over 12 weeks, three sessions per week, for a total of 36 sessions, with a preferred interval of approximately 48 hours between sessions. Each intervention-group session will last approximately 60-80 minutes and comprise 10 minutes of stationary cycling, one to two minutes of transition and preparation, 40-55 minutes of main exercise, and 5-10 minutes of cool-down, stretching, or manual techniques as indicated. At each session, approximately four patterns will be selected from the bank of 144 prespecified patterns, yielding 16–24 sets. Sets will generally comprise 8–12 repetitions or breathing cycles, with 60–120 s of rest, adjusted to preserve movement quality and the 40–55-min main exercise block. Detailed unilateral and bilateral dosing rules are provided in S4 File.

### Individualization and progression

Exercise variants will be selected using prespecified criteria for alignment, continuous breathing, control of the target region, pain, compensatory movement, and cue dependence. Progression requires at least eight correct repetitions or breathing cycles in every prescribed set; if these criteria are not met, the exercise will be modified or regressed. All decisions and their rationales will be documented. The complete selection, progression, regression, and manual-technique algorithm is provided in S4 File.

Progression will move from low-load positions to sitting, standing, weight-bearing, and functional tasks, with task-specific adaptations when relevant [12,13,28].

### Feedback

Feedback will initially be explicit and multimodal and will be progressively faded as performance improves; the complete feedback hierarchy is provided in S4 File [12,13,28].

### Recumbent stationary cycling

The warm-up will use a SEG BG-8229 recumbent stationary bicycle manufactured in Taiwan. In the initial sessions, the objectives will be to learn appropriate sitting posture, lower-limb alignment, and pedaling technique, followed by integration of breathing coordination. The bicycle has resistance settings from 1 to 20. The program will begin with alternating one minute at level 1 and one minute at level 2 for 10 minutes. The functional target is to cover 4 km in 10 minutes with appropriate technique, without overt compensation, and while retaining a perceived capacity to continue. Once this target is achieved consistently, the intervals will progress to levels 2 and 3 and subsequently to higher levels. Progression will be based on distance, the 0-10 Borg rating of perceived exertion, breathing, and movement quality rather than solely on the resistance setting.

### Stretching and manual techniques

Indication-based stretching and joint or soft-tissue techniques will be available to both groups. A static stretch will be held once per target muscle for 30 s when a restriction is present and for 20 s after the target range has been attained. When indicated, mobilization or soft-tissue techniques will be delivered as one set of 10 repetitions followed by immediate reassessment. The target, dose, response, and any adverse event will be documented; full procedures are provided in S4 File [29–31].

### Adherence and intervention fidelity

Attendance, exercises, sets, repetitions, resistance, modifications, feedback, manual techniques, and adverse events will be recorded at every session. Attendance adherence in both arms is defined as participation in at least 33 of the 36 sessions. This criterion reflects adherence to scheduled visits only and does not imply equal contact time or intervention dose between groups. Actual session duration, cumulative contact time with the provider, and the proportion of planned arm-specific components received will be recorded and reported separately. The per-protocol population will include participants who attend at least 33 sessions, receive at least 80% of the planned dose for their allocated arm, and have no major protocol deviation or prohibited concurrent intervention. Permitted, documented modifications made within the individualized framework will not be considered deviations. The threshold will be identical for women and men; menstrual symptoms may justify rescheduling or temporary modification. When a session is missed, a replacement time will be offered while preserving session order. Each month, a random 10% sample of intervention records will be audited by a researcher not directly involved in intervention delivery, and intervention details will be documented in accordance with TIDieR-Rehab and CERT [22,23].

### Active comparator and concomitant interventions

Participants in the active-comparator group will attend the same total of 36 sessions over 12 weeks and receive the general components shared by both groups: 10 minutes of stationary cycling, stretching of target muscles, prespecified mobilizations, and education on healthy movement habits. Each comparator session will last approximately 20-30 minutes. The type, sequence, dose, and delivery of shared components will be the same as the corresponding components in the intervention group. Education will address reducing prolonged time in static postures, increasing variability in daily movement, taking regular movement breaks, and applying basic postural principles during occupational and daily activities.

The active-comparator group will not receive the specific main component of the intervention: exercises based on the structured, staged model; retraining of muscle activation and coordination; training of basic and complex movement patterns; individualized progression of load and task; or the specific transition program toward health-related physical fitness. Attendance, actual contact time, components delivered, stretching and mobilization dose, educational content, and interventions received outside the study will be recorded in both groups.

The comparator was selected to control for the shared foundational components, number of visits, and exposure to the treatment environment, but it will not fully equalize provider-contact time or the volume of supervised activity. Equalizing time would have required sham activity, prolonged education, or general exercise whose absence of effects on posture, balance, physical activity, and participant expectations could not be guaranteed and could have changed the study question into a comparison of two exercise programs. The between-group contrast will therefore be interpreted as the added effect of the complete specific component of the systems-informed approach under pragmatic delivery conditions, not as the isolated effect of its exercise content independent of contact time, provider attention, or supervised-activity dose.

Participants in both groups will be asked to continue their usual activities and not begin a new structured exercise program. Sessions will be delivered individually or in groups of no more than three participants, with one provider for every three participants. Actual dose, durations of session components, completed sets, prolonged rest periods, and non-exercise interruptions will be recorded on the intervention-fidelity form.

### Outcomes and assessment schedule

The primary outcome is the vector of three postural components at week 12: the forward-head angle, forward-shoulder angle, and thoracic-kyphosis angle. No component alone will be treated as a surrogate for UCS. The primary comparison is the baseline-adjusted between-group difference in this vector. The same three variables will be measured at four-week follow-up to evaluate maintenance of change.

Secondary outcomes include active shoulder flexion and external rotation, active cervical flexion, EMG of selected scapular muscles, and static and dynamic lower-limb balance. EEG is exploratory and hypothesis-generating. Posture, range of motion, and balance will be recorded at baseline, week 12, and follow-up; EMG and EEG at baseline and week 12; and adverse events and adherence throughout the intervention.

### Measurement procedures

All assessments will follow written standard operating procedures, be performed by trained assessors, and occur at a similar time of day for each participant, within an allowable window of ±1 hour. Whenever possible, the same assessor will measure a given outcome at all time points for a participant. Participants will avoid strenuous activity for 24 hours and caffeine for three hours before electrophysiological assessments; sleep, medication, and recent activity will be recorded. Test order will remain fixed. Before enrollment of the first participant, intra-rater reliability for postural and range-of-motion measurements will be examined in a pilot sample. If the intraclass correlation coefficient is below 0.80 or measurement error exceeds the prespecified threshold, training and reliability testing will be repeated.

### Anthropometric and body-composition measurements

Standing height and body mass will be measured to the nearest 0.1 cm and 0.1 kg, respectively, using a Seca scale with an integrated stadiometer (Seca GmbH & Co. KG, Hamburg, Germany). Measurements will be obtained without shoes and while participants are wearing light clothing. Body mass index will be calculated as body mass in kilograms divided by height in metres squared (kg/m^2^). Body composition will be assessed using a BoCA X1 bioelectrical-impedance analyzer (Medigate Inc., Republic of Korea) in accordance with the manufacturer’s instructions. Body fat percentage and fat-free mass will be recorded descriptively and will not be treated as trial outcomes.

### Posture

Forward-head and forward-shoulder angles will be assessed using standardized lateral photogrammetry during screening and at baseline, week 12, and follow-up. Images will be acquired using a tripod-mounted Canon EOS M mirrorless digital camera (Canon Inc., Tokyo, Japan) fitted with a Canon EF-M 18–55 mm f/3.5–5.6 IS STM lens, set to a fixed focal length of 18 mm. Images will be recorded at a resolution of 5184 × 3456 pixels. The focal length, exposure settings, illumination, background, camera position, and participant position will be standardized during a pilot session and kept unchanged throughout the study.

The spinous process of C7 and the tragus and acromion on the dominant side will be marked. The camera’s optical axis will be perpendicular to the participant’s sagittal plane. The camera will be positioned 265 cm from the wall, the participant 23 cm from the wall, and the lens at shoulder height. A vertical reference and a calibration scale will be positioned in the participant’s sagittal plane. After three forward bends and three bilateral arm raises, the participant will stand comfortably with symmetrical weight distribution, without receiving any postural-correction cue, and three lateral images will be captured. The forward-head and forward-shoulder angles will be defined as the angles formed by the C7–tragus and C7– acromion lines, respectively, relative to the vertical reference. Angles will be measured using AutoCAD 2025, and the average of the three images will be used for analysis. The dominant side will be determined before baseline and used consistently across all assessments [7,24].

The flexible ruler will be used only to determine whether the kyphosis threshold is met at screening because it is practical, inexpensive, and has acceptable reliability and validity. After C7 and T12 have been marked, the thoracic curve will be traced onto paper. In the equation θ=4arctan(2H/L), L is the straight-line distance from C7 to T12, and H is the maximum perpendicular distance from the curve to line L. At baseline, week 12, and follow-up, the kyphotic angle (ICT-ITL) will be measured with DIERS formetric 4D and DICAM 3 software. The flexible ruler and Formetric serve different purposes, and their values will not be treated as directly interchangeable. Eligibility will be determined solely by screening; the Formetric result will not be used to reclassify participants. Longitudinal change will be assessed only with the same Formetric output. At each assessment, six scans will be acquired, with the participant stepping out of and then returning to the standardized position between scans, and their mean will be reported [32].

### Surface electromyography

Activity of the upper, middle, and lower trapezius and serratus anterior on the dominant side will be recorded using an eight-channel wireless Desktop DTS system (Noraxon USA Inc., Scottsdale, AZ, USA) and acquired using MyoResearch 3 (MR3) software. Four channels will record the target muscles, and a fifth channel placed over the middle deltoid will provide a temporal reference for movement onset. When required, the skin will be shaved, gently abraded, and cleaned with 70% alcohol. Disposable bipolar Ag/AgCl electrodes will be placed with a center-to-center distance of 20 mm, parallel to the muscle fibers and in accordance with SENIAM recommendations. Locations will be documented by measurement from anatomical landmarks and with a non-identifiable photograph [33,34].

Before recording, the resting signal and a brief contraction will be inspected. Raw signals will be processed using a fourth-order, zero-phase Butterworth band-pass filter from 20 to 450 Hz. A 50-Hz notch filter will be applied and documented only if persistent contamination remains after electrode connections have been corrected. After full-wave rectification, an RMS envelope will be calculated using a 50-ms moving window. Processing settings will remain constant across participants and time points.

For normalization, four maximum voluntary isometric contraction (MVIC) positions will be tested: a unilateral shoulder shrug in sitting; prone horizontal shoulder abduction with external rotation at 90 degrees of abduction; prone arm elevation along the line of the lower trapezius fibers at approximately 135 degrees of abduction; and resisted elevation in the scapular plane at 120-125 degrees. Each position will be performed three times for five seconds, with 60 seconds of rest between repetitions and two minutes between positions. Position order will be randomized, and resistance, stabilization, and instructions will be standardized. For each muscle, the highest stable one-second mean across all valid contractions will be used as the MVIC reference [33].

After five familiarization repetitions, participants will complete three recorded repetitions of active arm elevation in the scapular plane without external resistance and through their maximal pain-free range: a three-second raising phase, a one-second hold, and a three-second lowering phase, paced by a metronome, with 60 seconds of rest between repetitions. The terminal angle recorded at baseline will be reproduced at week 12. Synchronized video and an event marker will be used to verify timing and identify compensation. The principal amplitude metric will be the mean normalized RMS for the entire seven-second task, averaged across the three valid repetitions. Phase-specific values will be supportive analyses. The UT/MT, UT/LT, and UT/SA ratios will be calculated from the normalized amplitude over the full task.

For onset timing, the baseline mean and standard deviation will be calculated from the 500-ms window preceding movement onset. Muscle onset will be defined as the first point at which the signal envelope exceeds the baseline mean plus three standard deviations for at least 25 ms. Middle-deltoid onset will define time zero. A blinded analyst will verify detections using the raw signal, envelope, and synchronized video; if no reliable onset can be identified, the value will be treated as missing [7,35]. The normalized amplitudes of the four muscles and the three muscle ratios will form two principal EMG families. Phase-specific data and onset timing will be supportive analyses.

### Range of motion

Active flexion and external rotation of the dominant shoulder and active cervical flexion will be measured with a 360-degree universal goniometer with one-degree resolution [36]. For shoulder flexion, the participant will lie supine with the elbow extended and forearm neutral while the trunk and scapula are stabilized. The fulcrum will be positioned over the lateral aspect of the greater tubercle, the stationary arm parallel to the lateral midline of the thorax, and the moving arm along the humerus toward the lateral epicondyle.

Shoulder external rotation will be assessed in supine with the shoulder abducted to 90 degrees, the elbow flexed to 90 degrees, and the forearm neutral. A towel of standardized thickness will be placed beneath the humerus, and the scapula will be stabilized. The fulcrum will be positioned over the olecranon, the stationary arm perpendicular to the floor, and the moving arm along the dorsal surface of the ulna toward the ulnar styloid process.

Active cervical flexion will be assessed in sitting with the feet on the floor and the thoracic region in contact with the backrest. The fulcrum will be positioned over C7, the stationary arm vertical, and the moving arm along the line from C7 to the earlobe. The participant will bring the chin toward the chest without compensatory thoracic flexion. End range will be defined by the first compensatory movement, end-range resistance, or pain. Each movement will be performed three times with consistent instructions and speed, and the mean of the three measurements will be recorded [37].

### Static and dynamic balance

Static balance will be measured using DIERS pedoscan during the Romberg test with eyes open and eyes closed. The participant will stand barefoot with the feet in a comfortable, recorded position, arms by the sides, and gaze directed at a target at eye level. Three 35-second trials will be performed in each condition, with 60 seconds of rest between trials. The first five seconds will be discarded and the subsequent 30 seconds analyzed [38]. The mean of the three valid trials will be reported. Mean center-of-pressure velocity is the key measure; path length, sway area, and anteroposterior and mediolateral excursion are supportive measures.

Dynamic balance will be assessed using the Lower Quarter Y-Balance Test on the dominant limb. The dominant leg will be identified using the ball-kick test and will remain the assessed limb at every time point. Limb length will be measured from the anterior superior iliac spine to the medial malleolus. After six familiarization attempts in each direction, three valid trials will be recorded in the anterior, posteromedial, and posterolateral directions. Lifting the stance foot; leaning on or kicking the reach indicator; touching the floor with the reaching foot; or failing to return under control will invalidate the trial. Participants will rest for at least 10 seconds between trials and 30 seconds between directions. The greatest valid reach in each direction will be divided by limb length and multiplied by 100. The composite score will be the sum of the maximal reach distances in the three directions divided by three times limb length [39]. Results pertain only to performance of the dominant limb and will not be used to infer bilateral symmetry.

### Electroencephalography

EEG will be recorded using a portable EEG 3840 system (NR Sign [Negar Andishgan], Iran) with 24 channels and a sampling rate of 500 Hz. Nineteen scalp electrodes will be placed according to the international 10-20 system, with A1/A2 as references and horizontal and vertical electrooculography channels [40,41]. Electrode impedance will be maintained below 10 kΩ. After a five-minute adaptation period, recordings will be obtained during eyes-open sitting, eyes-closed sitting, eyes-open standing, and an eyes-open modified Matthiass test. Each condition will comprise three 60-second trials separated by 60 seconds of rest. Gaze instructions, target distance, foot and hand positions, and the procedure for marking the start and end of each trial will be standardized, and events will be marked in both the EEG file and synchronized video.

Preprocessing will be performed in EEGLAB 2026.0.0 using locked versions of clean_rawdata and ICLabel [41–43]. Flat-line segments will first be identified, followed by zero-phase band-pass filtering from 1 to 40 Hz. Bad-channel criteria will be a flat line longer than five seconds, correlation below 0.80 with a robust estimate based on the remaining channels, or line noise more than four standard deviations above the distribution. If more than 20% of scalp channels are classified as bad, that condition will be treated as missing. Otherwise, bad channels will be removed temporarily before independent component analysis (ICA), the data will be re-referenced to the average of the remaining channels, and the four conditions will be concatenated for extended Infomax ICA. Data rank will be calculated before ICA, and the number of components will be restricted to the effective rank. Components classified by ICLabel as ocular or muscular with a probability of at least 0.80 will be removed only after confirmation from their spatial pattern, time course, and EOG channels. Removed channels will subsequently be reconstructed by spherical interpolation, and all decisions will be documented.

The signal will be divided into non-overlapping two-second epochs. An epoch will be rejected if the amplitude of any channel exceeds ±100 μV or synchronized video shows overt movement. At least 60 acceptable epochs, equivalent to 120 seconds, will be required for each condition. Power spectral density will be calculated using Welch’s method with a one-second Hamming window and 50% overlap. Relative power will be calculated for delta (1 to <4 Hz), theta (4 to <8 Hz), alpha (8 to <13 Hz), and beta (13-30 Hz), relative to total power from 1 to 30 Hz. Regions will comprise frontal (F3/Fz/F4), central (C3/Cz/C4), and parietal (P3/Pz/P4) electrodes.

The primary exploratory EEG outcome is the change from baseline to week 12 in central relative alpha power during quiet standing with eyes open. Other conditions, regions, and frequency bands will be supplementary exploratory analyses. A sensitivity analysis will exclude recordings in which any central electrode was interpolated. The numbers of rejected channels, components, and epochs will be reported. Interpretation will focus on effect estimates, confidence intervals, and consistency of patterns; statistical significance alone will not be considered evidence of neural reorganization.

For the modified Matthiass test, the participant will stand barefoot with the feet parallel and hip-width apart, elevate both arms to 90 degrees of shoulder flexion with elbows extended and palms facing downward, and maintain the position for 60 seconds. Recording will begin after the arms reach 90 degrees. A step, displacement of either foot, contact with a support, or lowering of the arms will invalidate the trial, which will be repeated after rest [44].

### Sample-size justification

The joint covariance matrix of the three postural components and sufficient information on the correlations between baseline and week-12 values have not been published for this population. Consequently, the parameters required to calculate sample size directly for the planned MANCOVA on the basis of a prespecified clinically important effect were unavailable. The recruitment target was therefore determined before access to outcome data on the basis of resource constraints, operational feasibility, and the requirements for maintaining intervention fidelity.

The intervention is individualized, and during each session the provider must evaluate and document the quality of movement-pattern execution, compensatory movements, response to load, need for exercise selection, regression, or progression, and use of manual techniques for every participant. To preserve adequate opportunity for observation, feedback, and exercise modification, no more than three participants will be supervised concurrently by one provider, resulting in a provider-to-participant ratio of 1:3. With six delivery slots per day, a maximum of 18 participants can receive the intervention in groups of three over the course of a day. After randomization, each study arm will be assigned three non-consecutive days per week. The center can therefore provide all scheduled sessions to 36 participants, 18 per group, over the 12-week period.

Recruiting more participants would require increasing the number supervised concurrently, reducing the time devoted to observation and feedback, or increasing staffing and operating hours, any of which could weaken intervention fidelity. The fixed study period also precluded successive cohorts. Thus, the operational capacity of one cohort defined the prespecified ceiling, and the sample of 36 was not selected solely on the basis of sample sizes in earlier studies. Justification of sample size by resource constraints is methodologically acceptable when the study’s informational value and inferential limitations are reported transparently [45,46].

To describe the statistical sensitivity of this fixed sample size, an analysis based on the approximation of a two-group MANOVA with three dependent variables and an alpha level of 0.05 was conducted before access to outcome data. If evaluable data are available for all 36 participants, the design provides 80% power to detect a large multivariate effect of approximately *f^2^(V) = 0.34*, corresponding to Pillai’s *V ≈ 0.25.* If 29 evaluable participants remain after approximately 20% attrition or unusable data, the effect detectable with 80% power is approximately *f^2^(V) = 0.44*, corresponding to Pillai’s *V ≈ 0.31*. These are statistical sensitivity thresholds and should not be interpreted as expected effect sizes or thresholds of clinical importance. Because the covariance matrix and baseline-outcome correlations are unavailable, these calculations are only approximations for the planned MANCOVA.

The large effects reported by Seidi et al. are considered only contextual evidence that comprehensive corrective interventions may produce large responses. Owing to differences in the primary outcome and effect-size metric, those results were not used as the direct basis for the present sample size [7]. This study is not powered to detect small or moderate multivariate effects, perform subgroup analyses, or draw definitive conclusions regarding secondary outcomes or EEG. All results will be accompanied by effect estimates and 95% confidence intervals to show the range of values compatible with the observed data. Restricting the primary outcome to one omnibus multivariate test also addresses multiplicity in the primary analysis, but does not substitute for adequate statistical power. Effect estimates, the covariance structure, and their uncertainty from this study may inform more precise sample-size planning for subsequent trials.

### Data management and quality assurance

Each participant will receive a non-identifying study code. The identity-linkage file will be stored separately in encrypted form with restricted access. Paper forms will be stored in a locked cabinet, and electronic files in encrypted university storage. Key variables will be entered independently twice, and discrepancies will be resolved against source documents. Range checks, missing data, timestamps, and file modifications will be documented in an audit trail. A locked database version will be created before the group code is broken. Data will be retained for at least five years or for the longer period required by university regulations.

### Statistical analysis

Analyses will be conducted in R according to the intention-to-treat principle. The R version, statistical packages, and analysis code will be documented and archived in the statistical analysis plan before the group code is broken. Baseline characteristics will be summarized using appropriate descriptive statistics, without significance testing for baseline between-group differences. All tests will be two-sided, and alpha for the primary test will be 0.05. Sex will be included in the models because it is used for stratified randomization; no subgroup analysis or intervention-by-sex interaction test will be performed.

The primary estimand is the adjusted difference between the systems-informed intervention group and the active-comparator group in the three-variable postural profile at week 12. This contrast estimates the added effect of the specific staged component of the systems-informed intervention beyond the shared components of stationary cycling, stretching, mobilization, and general education on movement habits. The three week-12 angles will be entered as dependent variables in a MANCOVA, with group and sex as fixed factors and the baseline values of the same three angles as covariates. The omnibus test will use Pillai’s trace. A significant omnibus test does not by itself indicate improvement in all three components. Only if the omnibus test is significant will three outcome-specific ANCOVAs be performed with the same adjustment variables, with P values corrected using the Holm method. Concurrent improvement will be claimed only if all three effect estimates favor correction and all three component tests remain significant after Holm correction. If assumptions are seriously violated, a permutation-based multivariate analysis that preserves the covariates and restrictions arising from stratified randomization will be performed as a sensitivity analysis.

Longitudinal analyses of posture, range of motion, and balance will use constrained linear mixed models. Baseline, week 12, and follow-up measurements will be entered as repeated responses. Categorical time, sex, and group-by-time interactions for week 12 and follow-up will be fixed effects; omission of the group main effect will constrain baseline group means to equality. A random intercept will be included for each participant. Models will be estimated using restricted maximum likelihood, with Satterthwaite degrees of freedom. Prespecified contrasts are the between-group differences at week 12 and follow-up. Week-12 postural contrasts from these models are supportive of the primary analysis; primary component-level inference will follow the hierarchical MANCOVA-ANCOVA procedure. The three postural contrasts at follow-up will constitute one family and will be Holm-adjusted. At each post-baseline time point, the three range-of-motion outcomes will form one family, and mean center-of-pressure velocity and the Y-Balance composite score will form a second family; Holm correction will be applied within each family. Other center-of-pressure measures and direction-specific Y-Balance scores will be supportive analyses.

Week-12 EMG outcomes will be compared using ANCOVA adjusted for baseline values and sex. The normalized amplitudes of the four muscles and the three muscle ratios will form two separate families, with Holm correction applied within each family. Skewed ratios will be transformed using the natural logarithm, and results will be back-transformed and reported as ratios of geometric means with 95% confidence intervals. Phase-specific values and onset timing are supportive analyses. The primary exploratory EEG outcome will be analyzed using ANCOVA adjusted for baseline values and sex. All other condition-by-region-by-band combinations will form one exploratory family, for which the false-discovery rate will be controlled at 5% using the Benjamini-Hochberg procedure.

For individual outcomes, adjusted mean differences, 95% confidence intervals, and Hedges’ g will be reported. Hedges’ g will be calculated from the adjusted between-group difference and pooled baseline standard deviation. If primary-outcome data are missing, multiple imputations by chained equations will be performed with at least 50 imputed datasets. The imputation model will include group, sex, postural values at baseline and post-baseline time points, and a parsimonious set of prespecified auxiliary variables. Predictive mean matching will be used for continuous variables. In each imputed dataset, a full multivariate model and a reduced model without the group effect will be fitted, and the omnibus group effect will be pooled using a multivariate Wald test with the D1 method. Estimates for individual outcomes will be pooled using Rubin’s rules. Pillai’s V will be reported in the complete-case analysis.

Complete-case and per-protocol analyses will be presented as sensitivity analyses. The per-protocol population will comprise participants who attend at least 33 of 36 sessions, receive at least 80% of the planned dose for their assigned arm, and have no major protocol deviation, including initiation of a structured exercise program outside the study. Permitted and documented modifications within the individualized intervention rules will not be considered protocol deviations. Because session duration and content differ between the two arms, dose adherence will be calculated relative to the planned program for the assigned arm; cumulative contact time, actual session duration, and percentage of planned dose received will also be reported separately by group. If more than 10% of primary-outcome data are missing, a sensitivity analysis incorporating departures from the missing-at-random assumption will be performed. Last observation carried forward and an interim efficacy analysis will not be used.

### Safety, adverse events, and insurance

Before each session, participants will be asked about pain, new injury, and changes in medical status. Potential adverse events include transient muscle soreness, subacute symptom aggravation, tenderness or bruising after soft-tissue release, and abdominal discomfort from contact with the exercise ball. Pain intensity will be recorded on a 0-10 scale. Pain greater than 5 or pain that prevents correct execution will prompt cessation of the exercise, assessment of load, range, and involved tissue, and appropriate modification. Persistent pain, neurological symptoms, unusual dyspnea, dizziness, or another warning sign will result in termination of the session and medical referral. The intervention will be discontinued when medically indicated or when risk persists, but a single report of pain will not by itself lead to exclusion from the study. All adverse events will be documented with their severity, possible relationship to the intervention, and outcome [22].

Sports insurance will remain valid for all participants from before formal assessment until completion of follow-up, and the intervention center holds public-liability insurance. Any incident will be reported within the required period. Reimbursement will be governed by the terms of the insurance policy, and complete coverage of all expenses cannot be guaranteed.

### Trial monitoring and stopping rules

Given the low-risk intervention, limited sample size, short study duration, and absence of an interim efficacy analysis, a formal data monitoring committee will not be established. An independent trial-safety and conduct monitor will nevertheless be appointed before recruitment begins. This person will have no role in intervention design, recruitment, exercise delivery, outcome assessment, data analysis, or manuscript preparation, and will have no financial, employment, or professional dependency relationship with the principal investigator, the intervention clinic, or the funder.

The independent monitor will review coded reports on recruitment and informed consent, adverse events, withdrawals, medical referrals, major protocol deviations, and completeness of safety records after the first six participants have enrolled and monthly thereafter. Efficacy results will not be made available to the monitor, and group allocation will be revealed only if required to evaluate the safety of an adverse event. Any serious adverse event potentially related to the intervention will be reported to the monitor within 24 hours, and recruitment and intervention delivery will be suspended temporarily pending initial review. Two unexpected moderate or severe adverse events with a similar pattern and a possible relationship to one intervention component will likewise result in temporary suspension and review of that component.

The independent monitor may recommend temporary suspension of recruitment or intervention delivery, protocol modification, medical referral, or immediate reporting to the ethics committee. A temporary safety suspension will be implemented immediately. Resumption, modification, or termination of the study will be decided after the monitor’s review and ethics-committee approval. Monitor reports will be sent directly to the ethics committee and research team and archived until study completion. The study may also be suspended because of major disruption in access to the center, security conditions, prolonged equipment failure, or an ethics-committee directive.

### Ethical principles, confidentiality, and incidental findings

The study will be conducted in accordance with the 2024 Declaration of Helsinki and the principles of ICH E6(R3). Participation is voluntary, and participants may withdraw without adverse consequences. If a participant discontinues exercise, outcome follow-up will continue with permission. Requests to remove data will be honored until the data have been de-identified and the database locked; after that point, it may not be possible to separate some aggregated data.

EEG in this study is not a diagnostic procedure. If the operator or neurologist suspects a potentially important finding, it will first be reviewed by the neurologist. If it is considered clinically important, the participant will be informed confidentially and referred for appropriate evaluation. Identifiable images and videos will not be included in any public dataset.

### Protocol access, data sharing, and dissemination

The approved protocol, statistical analysis plan, data dictionary, analysis code, and the de-identified individual-level minimal dataset will be deposited in a stable public repository when the trial results are published. Sensitive raw data, including videos, identifiable images, and raw signals that carry a risk of re-identification, will not be made publicly available. Research access to these materials will require a justified request, approval by the relevant institutional body or ethics committee, and a data-use agreement. Access decisions will not be controlled by a single author. Results will be published regardless of their direction or statistical significance, and a plain-language summary will be made available to participants.

Any important future protocol modification will be submitted to the Research Ethics Committee, updated in the trial registry, communicated to the investigators and participants when relevant, and described in the final trial report.

Trial results will be updated in the trial registry, submitted for peer-reviewed publication, presented to relevant academic and clinical audiences, and summarized in plain language for participants and the public.

### Patient and public involvement

No patients, prospective participants, or members of the public were involved in formulating the research question, designing the trial, selecting the outcomes, developing the intervention, or preparing the protocol. Their role is limited to participation in the trial. After completion of the study, a plain-language summary of the findings will be made available to participants.

### Use of artificial intelligence tools

ChatGPT (OpenAI) was used during preparation of this protocol to assist with manuscript organization, Persian-to-English translation, language editing, and checking alignment with reporting guidelines. The tool was not used to generate or analyze participant data. All AI-assisted text was critically reviewed and revised by the authors, and all references and methodological statements were checked against the original sources and applicable reporting guidance. All scientific and methodological decisions were made by the authors, who take full responsibility for the accuracy, originality, and integrity of the manuscript and its supporting files. No participant-level data or confidential study records were entered into the tool.

## Discussion

Available evidence indicates that corrective exercise can improve postural alignment in people with UCS, but heterogeneity in eligibility criteria, intervention components, and exercise-progression rules limits interpretation and reproducibility [2–5]. By requiring the concurrent presence of three postural components and defining a multivariate primary outcome, the present study will evaluate the response of the combined pattern. This phenotype is an operational research definition and not a validated diagnostic criterion for UCS.

The program’s innovation does not lie in a single new exercise, but in organizing established components within the criterion-based staged model, applying explicit selection and progression rules, and documenting provider decisions. This structure permits individualization within a shared framework and is consistent with recommendations for reporting complex and rehabilitation interventions [22,23,47,48].

EMG, range of motion, and balance are assessed to describe whether postural change is accompanied by neuromuscular and functional responses, not to prove a mechanism. EEG has likewise been restricted to one primary exploratory outcome; a positive finding will therefore be hypothesis-generating and will not be sufficient to claim neural reorganization.

### Strengths and limitations

A distinctive feature of this study is that corrective exercise is treated as one stage within a clinical continuum rather than as an intervention that ends when postural angles change. Restoration of range of motion, retraining of muscle activation, and control of movement patterns are intended to provide a foundation for progression to load tolerance, health-related physical fitness, self-management, and more sustained participation in physical activity. This approach assumes that physical inactivity, prolonged static postures, and repeated exposure to monotonous movement patterns may contribute to the persistence of unfavorable movement strategies [49]. The present study tests this pathway but does not evaluate long-term prevention of recurrence.

Explicit rules for exercise selection, regression, and progression; documentation of provider decisions; intervention-fidelity audits; and criteria for transition to health-enhancing physical activity bring the theoretical model closer to an implementable, reproducible clinical pathway and may reduce the gap between controlled research protocols and individualized decision-making in clinical practice. Nevertheless, this study is not a definitive clinical guideline. The four-week follow-up evaluates only short-term maintenance of change and the feasibility of transition to self-management.

Methodological strengths include the prespecified phenotype definition, multivariate primary outcome, stratified randomization, allocation concealment, blinding of assessors and analysts, objective measurement tools, and explicit separation of confirmatory, secondary, and exploratory analyses. Conversely, participants and the intervention provider cannot be blinded; the small sample is expected to be adequate only for large omnibus effects; and the single-center setting, age range of 18-40 years, and restriction to participants with BMI in the normal range limit generalizability.

Use of an active comparator permits estimation of the added effect of the specific progressive component beyond stationary cycling, stretching, mobilization, and general education, and provides a more conservative test than comparison with no intervention. However, provider-contact time and the volume of supervised exercise are greater in the intervention group. Between-group differences may therefore reflect the combined effects of the specific intervention content, a greater exercise dose, provider attention, individualized feedback, and participant expectations; the independent contributions of these factors cannot be determined. Blinding of assessors and analysts, objective outcomes, and documentation of actual contact time may reduce some sources of bias but do not eliminate this design limitation. Findings should therefore not be interpreted as the specific effect of the systems-informed approach independent of time and provider attention, or as the effect of the entire package compared with no treatment. Separating these effects will require future trials with time-and attention-matched control conditions or designs that dismantle intervention components.

Individualization is both a strength and a limitation. Although common rules govern the intervention, progression pathways and the use of manual techniques may differ among participants. The trial will therefore evaluate the effectiveness of the complete framework but will not determine the independent contribution of any exercise or manual technique.

In summary, this trial will determine whether adding a 12-week structured and staged program to shared cycling, stretching, mobilization, and education produces greater improvement in the postural profile and whether any change is accompanied by neuromuscular responses, range-of-motion changes, and balance changes. Regardless of the direction of the findings, transparent reporting of decision rules and actual dose will support replication and critical appraisal of this approach.

## Data Availability Statement

No datasets were generated or analyzed for this protocol article. Data-sharing arrangements for the completed trial, including controlled-access procedures for potentially identifiable videos, images, and raw biosignal recordings, are described in the Methods.

## Author contributions

Mohamad Khorami Moghadam: Conceptualization, Methodology, Investigation, Intervention development, Project administration, Visualization, and Writing – original draft. Gholam Ali Ghasemi: Conceptualization, Methodology, Supervision, and Writing – review and editing. Leila PoorSaadat: EEG methodology and supervision and Writing – review and editing. Masoud Golpayegani: EMG and postural-assessment methodology and supervision and Writing – review and editing. All authors reviewed and approved the final manuscript and accepted responsibility for the work.

## Acknowledgments

The authors thank the staff of the Sport Sciences Laboratory at Arak University and Valiasr Hospital in Arak for their assistance with study preparation.

## Supporting information

**S1** Checklist. Completed SPIRIT 2025 checklist.

**S2** File. Ethics-approved study protocol in the original Persian language.

**S3** File. English translation of the ethics-approved study protocol.

**S4** File. Supplementary intervention manual, including the decision matrix, exercise-pattern bank, and rules for exercise selection, regression, progression, dose, and fidelity recording.

**S5** File. Final locked statistical analysis plan.

## References

1. Singla D, Veqar Z. Association Between Forward Head, Rounded Shoulders, and Increased Thoracic Kyphosis: A Review of the Literature. J Chiropr Med. 2017;16: 220–229. doi:10.1016/j.jcm.2017.03.004

2. Sepehri S, Sheikhhoseini R, Piri H, Sayyadi P. The effect of various therapeutic exercises on forward head posture, rounded shoulder, and hyperkyphosis among people with upper crossed syndrome: a systematic review and meta-analysis. BMC Musculoskelet Disord. 2024;25: 105. doi:10.1186/s12891-024-07224-4

3. Khorramroo F, Rostami M, Bafrouei MJ. Corrective exercises strongly improve posture but fail to produce consistent clinical or functional benefits in patients with upper crossed syndrome: a systematic review and meta-analysis of randomized controlled trials. BMC Sports Sci Med Rehabil. 2026;18: 302. doi:10.1186/s13102-026-01707-8

4. Chaudhuri S, Chawla JK, Phadke V. Physiotherapeutic Interventions for Upper Cross Syndrome: A Systematic Review and Meta-Analysis. Cureus. 2023;15: e45471. doi:10.7759/cureus.45471

5. Chang MC, Choo YJ, Hong K, Boudier-Revéret M, Yang S. Treatment of Upper Crossed Syndrome: A Narrative Systematic Review. Healthcare. 2023;11: 2328. doi:10.3390/healthcare11162328

6. Yaghoubitajani Z, Gheitasi M, Bayattork M, Andersen LL. Corrective exercises administered online vs at the workplace for pain and function in the office workers with upper crossed syndrome: randomized controlled trial. Int Arch Occup Environ Health. 2022;95: 1703–1718. doi:10.1007/s00420-022-01859-3

7. Seidi F, Bayattork M, Minoonejad H, Andersen LL, Page P. Comprehensive corrective exercise program improves alignment, muscle activation and movement pattern of men with upper crossed syndrome: randomized controlled trial. Sci Rep. 2020;10: 20688. doi:10.1038/s41598-020-77571-4

8. Bayattork M, Seidi F, Minoonejad H, Andersen LL, Page P. The effectiveness of a comprehensive corrective exercises program and subsequent detraining on alignment, muscle activation, and movement pattern in men with upper crossed syndrome: protocol for a parallel-group randomized controlled trial. Trials. 2020;21: 255. doi:10.1186/s13063-020-4159-9

9. Ivanenko Y, Gurfinkel VS. Human Postural Control. Front Neurosci. 2018;12: 171. doi:10.3389/fnins.2018.00171

10. Jeong GH, Lee BH. Effects of Telerehabilitation Combining Diaphragmatic Breathing Re-Education and Shoulder Stabilization Exercises on Neck Pain, Posture, and Function in Young Adult Men with Upper Crossed Syndrome: A Randomized Controlled Trial. J Clin Med. 2024;13: 1612. doi:10.3390/jcm13061612

11. Sannasi R, Dakshinamurthy A, Dommerholt J, Desai V, Kumar A, Sugavanam T. Diaphragm and core stabilization exercises in low back pain: A narrative review. J Bodyw Mov Ther. 2023;36: 221–227. doi:10.1016/j.jbmt.2023.07.008

12. Gokeler A, Neuhaus D, Benjaminse A, Grooms DR, Baumeister J. Principles of Motor Learning to Support Neuroplasticity After ACL Injury: Implications for Optimizing Performance and Reducing Risk of Second ACL Injury. Sports Med. 2019;49: 853–865. doi:10.1007/s40279-019-01058-0

13. Chua L-K, Jimenez-Diaz J, Lewthwaite R, Kim T, Wulf G. Superiority of external attentional focus for motor performance and learning: Systematic reviews and meta-analyses. Psychol Bull. 2021;147: 618–645. doi:10.1037/bul0000335

14. Rubega M, Formaggio E, Di Marco R, Bertuccelli M, Tortora S, Menegatti E, et al. Cortical correlates in upright dynamic and static balance in the elderly. Sci Rep. 2021;11: 14132. doi:10.1038/s41598-021-93556-3

15. Bakker LBM, Lamoth CJC, Vetrovsky T, Gruber M, Caljouw SR, Nieboer W, et al. Neural Correlates of Balance Skill Learning in Young and Older Individuals: A Systematic Review and Meta-analysis. Sports Med Open. 2024;10: 3. doi:10.1186/s40798-023-00668-3

16. Formaggio E, Bertuccelli M, Rubega M, Marco R Di, Cantele F, Gottardello F, et al. Brain oscillatory activity in adolescent idiopathic scoliosis. Sci Rep. 2022;12: 17266. doi:10.1038/s41598-022-19449-1

17. Jung J-Y, Lee Y-B, Kang C-K. Effect of Forward Head Posture on Resting State Brain Function. Healthcare. 2024;12: 1162. doi:10.3390/healthcare12121162

18. Jung J-Y, Kang C-K. Effect on Discomfort and Attention Through Analysis of Resting-State Brain Wave Activity in Forward Head Posture. Korean Society for Emotion and Sensibility. 2024;27: 105–112. doi:10.14695/KJSOS.2024.27.2.105

19. Chan A-W, Boutron I, Hopewell S, Moher D, Schulz KF, Collins GS, et al. SPIRIT 2025 statement: Updated guideline for protocols of randomised trials. PLoS Med. 2025;22: e1004589. doi:10.1371/journal.pmed.1004589

20. Hróbjartsson A, Boutron I, Hopewell S, Moher D, Schulz KF, Collins GS, et al. SPIRIT 2025 explanation and elaboration: updated guideline for protocols of randomised trials. BMJ. 2025;389: e081660. doi:10.1136/bmj-2024-081660

21. Hopewell S, Chan A-W, Collins GS, Hróbjartsson A, Moher D, Schulz KF, et al. CONSORT 2025 statement: updated guideline for reporting randomised trials. BMJ. 2025;389: e081123. doi:10.1136/bmj-2024-081123

22. Signal N, Olsen S, Gomes E, McGeorge C, Taylor D, Alder G. Developing the TIDieR-Rehab checklist: a modified Delphi process to extend the Template for Intervention Description and Replication (TIDieR) for rehabilitation intervention reporting. BMJ Open. 2024;14: e084319. doi:10.1136/bmjopen-2024-084319

23. Slade SC, Dionne CE, Underwood M, Buchbinder R. Consensus on Exercise Reporting Template (CERT): Explanation and Elaboration Statement. Br J Sports Med. 2016;50: 1428–1437. doi:10.1136/bjsports-2016-096651

24. Mylonas K, Tsekoura M, Billis E, Aggelopoulos P, Tsepis E, Fousekis K. Reliability and Validity of Non-radiographic Methods of Forward Head Posture Measurement: A Systematic Review. Cureus. 2022;14: e27696. doi:10.7759/cureus.27696

25. Gadotti IC, Armijo-Olivo S, Silveira A, Magee D. Reliability of the Craniocervical Posture Assessment: Visual and Angular Measurements Using Photographs and Radiographs. J Manipulative Physiol Ther. 2013;36: 619–625. doi:10.1016/j.jmpt.2013.09.002

26. Abarca-Gómez L, Abdeen ZA, Hamid ZA, Abu-Rmeileh NM, Acosta-Cazares B, Acuin C, et al. Worldwide trends in body-mass index, underweight, overweight, and obesity from 1975 to 2016: a pooled analysis of 2416 population-based measurement studies in 128·9 million children, adolescents, and adults. The Lancet. 2017;390: 2627–2642. doi:10.1016/S0140-6736(17)32129-3

27. Kuiken TA, Lowery MM, Stoykov NS. The effect of subcutaneous fat on myoelectric signal amplitude and cross-talk. Prosthet Orthot Int. 2003;27: 48–54. doi:10.3109/03093640309167976

28. Aoyagi Y, Ohnishi E, Yamamoto Y, Kado N, Suzuki T, Ohnishi H, et al. Feedback protocol of “fading knowledge of results” is effective for prolonging motor learning retention. J Phys Ther Sci. 2019;31: 687–691. doi:10.1589/jpts.31.687

29. Ingram LA, Tomkinson GR, d’Unienville NMA, Gower B, Gleadhill S, Boyle T, et al. Optimising the Dose of Static Stretching to Improve Flexibility: A Systematic Review, Meta-analysis and Multivariate Meta-regression. Sports Medicine. 2025;55: 597–617. doi:10.1007/s40279-024-02143-9

30. Wang S, Zeng J, Chapple CM, Mani R, Ribeiro DC. Initial effect of high-volume mobilisation with movement on shoulder range of motion and pain in patients with rotator cuff-related shoulder pain: protocol for a randomised controlled trial (Evolution Trial). BMJ Open. 2023;13: e069919. doi:10.1136/bmjopen-2022-069919

31. Wang S, Chapple C, Farrell G, Quinn D, Tumilty S, Ribeiro DC. Dosage of joint mobilization for the management of patients with rotator cuff-related shoulder pain: A scoping review. Musculoskelet Sci Pract. 2024;70: 102903. doi:10.1016/j.msksp.2023.102903

32. Degenhardt BF, Starks Z, Bhatia S. Reliability of the DIERS Formetric 4D Spine Shape Parameters in Adults without Postural Deformities. Biomed Res Int. 2020;2020: 1796247. doi:10.1155/2020/1796247

33. Ekstrom RA, Soderberg GL, Donatelli RA. Normalization procedures using maximum voluntary isometric contractions for the serratus anterior and trapezius muscles during surface EMG analysis. Journal of Electromyography and Kinesiology. 2005;15: 418–428. doi:10.1016/j.jelekin.2004.09.006

34. Hermens HJ, Freriks B, Disselhorst-Klug C, Rau G. Development of recommendations for SEMG sensors and sensor placement procedures. Journal of Electromyography and Kinesiology. 2000;10: 361–374. doi:10.1016/S1050-6411(00)00027-4

35. Mendez-Rebolledo G, Gatica-Rojas V, Martinez-Valdes E, Xie HB. The recruitment order of scapular muscles depends on the characteristics of the postural task. Journal of Electromyography and Kinesiology. 2016;31: 40–47. doi:10.1016/j.jelekin.2016.09.001

36. Pérez-de la Cruz S, de León ÓA, Mallada NP, Rodríguez AV. Validity and intra-examiner reliability of the Hawk goniometer versus the universal goniometer for the measurement of range of motion of the glenohumeral joint. Med Eng Phys. 2021;89: 7–11. doi:10.1016/j.medengphy.2021.01.005

37. Araujo GGC, Pontes-Silva A, Leal P da C, Gomes BS, Reis ML, de Mello Pereira Lima SK, et al. Goniometry and fleximetry measurements to assess cervical range of motion in individuals with chronic neck pain: a validity and reliability study. BMC Musculoskelet Disord. 2024;25: 651. doi:10.1186/s12891-024-07775-6

38. Koltermann JJ, Gerber M, Beck H, Beck M. Validation of Different Filters for Center of Pressure Measurements by a Cross-Section Study. Technologies (Basel). 2019;7: 68. doi:10.3390/technologies7040068

39. Plisky P, Schwartkopf-Phifer K, Huebner B, Garner MB, Bullock G. Systematic Review and Meta-Analysis of the Y-Balance Test Lower Quarter: Reliability, Discriminant Validity, and Predictive Validity. Int J Sports Phys Ther. 2021;16: 1190–1209. doi:10.26603/001c.27634

40. Seeck M, Koessler L, Bast T, Leijten F, Michel C, Baumgartner C, et al. The standardized EEG electrode array of the IFCN. Clinical Neurophysiology. 2017;128: 2070–2077. doi:10.1016/j.clinph.2017.06.254

41. Pernet C, Garrido MI, Gramfort A, Maurits N, Michel CM, Pang E, et al. Issues and recommendations from the OHBM COBIDAS MEEG committee for reproducible EEG and MEG research. Nat Neurosci. 2020;23: 1473–1483. doi:10.1038/s41593-020-00709-0

42. Pion-Tonachini L, Kreutz-Delgado K, Makeig S. ICLabel: An automated electroencephalographic independent component classifier, dataset, and website. Neuroimage. 2019;198: 181–197. doi:10.1016/j.neuroimage.2019.05.026

43. Delorme A, Makeig S. EEGLAB: an open source toolbox for analysis of single-trial EEG dynamics including independent component analysis. J Neurosci Methods. 2004;134: 9–21. doi:10.1016/j.jneumeth.2003.10.009

44. Albertsen IM, Dettmann K, Babin K, Stücker R, Schröder J, Zech A, et al. Spinal postural changes during the modified Matthiass test in healthy children. Orthopade. 2018;47: 567–573. doi:10.1007/s00132-018-3558-z

45. Lakens D. Sample Size Justification. Collabra Psychol. 2022;8: 33267. doi:10.1525/collabra.33267

46. Giner-Sorolla R, Montoya AK, Reifman A, Carpenter T, Lewis NA, Aberson CL, et al. Power to Detect What? Considerations for Planning and Evaluating Sample Size. Personality and Social Psychology Review. 2024;28: 276–301. doi:10.1177/10888683241228328

47. Hoffmann TC, Glasziou PP, Boutron I, Milne R, Perera R, Moher D, et al. Better reporting of interventions: template for intervention description and replication (TIDieR) checklist and guide. BMJ. 2014;348: g1687. doi:10.1136/bmj.g1687

48. Skivington K, Matthews L, Simpson SA, Craig P, Baird J, Blazeby JM, et al. A new framework for developing and evaluating complex interventions: update of Medical Research Council guidance. BMJ. 2021;374: n2061. doi:10.1136/bmj.n2061

49. Bull FC, Al-Ansari SS, Biddle S, Borodulin K, Buman MP, Cardon G, et al. World Health Organization 2020 guidelines on physical activity and sedentary behaviour. Br J Sports Med. 2020;54: 1451–1462. doi:10.1136/bjsports-2020-102955

